# Consent is a confounding factor in a prospective observational study of critically ill elderly patients

**DOI:** 10.1101/2022.03.18.22272593

**Authors:** Hans Flaatten, Bertrand Guidet, Christian Jung, Ariane Boumendil, Susannah Leaver, Wojciech Szczeklik, Antonio Artigas, Finn Andersen, Rui Moreno, Sten Walther, Sandra Oeyen, Joerg C. Schefold, Brian Marsh, Michael Joannidis, Muhammed Elhadi, Yuriy Nalapko, Jesper Fjølner, Dylan W. de Lange

## Abstract

During analysis of a prospective multinational observation study of critically ill patients ≥80 years of age, the VIP2 study, we also studied the effects of differences in country consent for study inclusion. This is a post hoc analysis where the ICUs were analyzed according to requirement for study consent. Group A: ICUs in countries with no requirement for consent at admission but with deferred consent in survivors. Group B: ICUs where some form of active consent at admission was necessary either from the patient or surrogates. Patients’ characteristics, the severity of disease and outcome variables were compared. Totally 3098 patients were included from 21 countries. The median age was 84 years (IQR 81-87). England was not included because of changing criteria for consent during the study period. Group A (7 countries, 1220 patients), and group B (15 countries, 1878 patients) were comparable with age distribution, organ failure score, frailty score and activity of daily life. Cognition was better preserved prior to admission in group B. Group A suffered from more organ dysfunction at admission compared to group B with Sequential Organ Failure Assessment score median 8 and 5 respectively. Survival at 30-day was lower in group A, 50.8 % compared to 66.7% in group B (p<0.001). We found profound effects on outcomes according to differences in obtaining consent for this study. It seems that the most severely ill elderly patients were less often recruited to the study in group B. Hence the outcome measured as survival was higher in this group.

We therefore conclude that consent likely is an important confounding factor for outcome evaluation in international studies focusing on old patients.

## Introduction

In a recently published, prospective, observational study of the acutely admitted very old intensive care patients (≥ 80 years old), the VIP2 study) conducted in 22 European countries, we found an overall 30-day survival of 61%. Factors predicting mortality were frailty at admission, ICU admission categories and degree of organ failure at admission [1]. During the study, we also noticed a considerable heterogeneity among European countries regarding the requirements for informed consent for this observational study. Some countries waived the need for informed consent while others demanded consent prior to patient inclusion.

Previously we have reported problems with obtaining consent for a similar study with waiting time before decision up to one year for some countries [2].

In observational clinical research, the patients are typically not subjected to an intervention that can alter the course of the disease or illness. Only patient data that is already collected in the daily clinical routine or collected for the purpose of that prospective observational research, are used. In general, patients should have the right to decide whether they want their data to enter a research project, also in non-intervention studies. However, informed consent from patients is not always possible, particularly in acute care settings. When a patient is severely ill and admitted to an intensive care unit (ICU), they very often lack decisional capacity. In such instances, the national or regional medical ethical committee might agree to use informed consent from surrogate decision-makers or waive the need for informed consent at the time of admission, so-called deferred consent, and inform patient survivors about the study and their right to withdraw their inclusion at a later stage. The introduction of the General Data Protection Act (GDPR) in the European Union (EU) May 2018 necessitated in many countries an additional (written) informed consent for sharing privacy sensitive patent data even when deferred consent was allowed by the ethical committees. We started recruiting countries and ICUs to the VIP2 study in spring 2018, just before to the implementation of the European General Data Protection Regulation (GDPR). This resulted that some countries waived the need for informed consent while others, especially those who completed the ethical procedures after implementation of the GDPR, required written informed consent from either the critically ill patient themselves, or, if not possible, from next-of-kin.

During the preparation and analysis of the main publication from the VIP2 study, we were concerned about any effect from these two different approaches regarding consent or no consent with potential bias on the recruitment to the study.

In this paper, our aim was to reveal any effect from consent on patient demographics, in particular about presence of organ failure at admission and later mortality of the enrolled patients.

## Materials and Methods

The methods are shortly described below, and a detailed overview can be found in the recent publication from the study [1]. The main aim of the VIP2 study was to investigate the influence of common geriatric syndromes like frailty, cognitive decline, activity of daily life and comorbidity/polypharmacy on various outcome measures, mainly ICU resource use and mortality. The severity of the critical illness was measured using the sequential organ dysfunction score (SOFA) [2] at admission.

To analyze potential effects from different requirements for ethical approval, we divided the countries into two groups. Those allowed to include patients without consent at ICU admission (group A), although most required deferred consent in hospital survivors; and countries that needed patient or legal proxy consent at admission, be it from the patient, caregivers or independent physicians (Group B). To illustrate the effects, we analyzed the presence of geriatric syndromes clinical frailty scale (CFS) and Informant Questionnaire on Cognitive Decline in the Elderly (IQCODE) and SOFA score at admission. For outcomes, we used ICU length of stay (LOS), use of intensive care procedures for organ support and 30-day survival. The groups were compared using confidence intervals of the appropriate variables. For proportions (outcome) Chi-square test was also applied. (SPSS v 25). Regression analysis including the above-mentioned variables and consent/no consent as a dichotomousvariable was also performed.

The main study was registered at ClinicalTrials.gov (ID: NCT03370692), and national or regional regulatory approval were given.

## Results

The main study recruited 3920 patients with a median age of 84 years (interquartile range, IQR, from 81-87) from 242 ICUs in 22 countries. The overall 30-day survival was 61.2%. The median duration of patient recruitment in an ICU was 64 days with no differences in outcomes in the ICUs below or above the median duration of recruitment (data not shown).

One country, the United Kingdom, initially required full consent but later accepted inclusion without consent for patients who died prior to consent, leaving 3098 in this post-hoc study. Hence, this country was excluded from the main analysis but included in a sensitivity analysis since they were the largest contributing country with 822 recruited patients.

Another country, the Netherlands, initially approved to include patients without consent, but after the introduction of GDPR, 5 hospitals changed this to informed consent. Hence this country appears with ICUs in both groups. The group A consisted of 7 countries with 1220 patients included and group B of 15 countries with 1878 patients. Table 1 (Table 1 here) shows the baseline characteristics at admission in the two groups.

**Table 1.** Admission data in all included patients.

|  | Group A (no consent) | Group B (consent) |
| --- | --- | --- |
| Countries (n) | 7 <sup>c</sup> | 15 <sup>d</sup> |
| Patients analysed | 1220 | 1878 |
| Missing outcome data | 45 (3.7%) | 25 (1.3%) |
| Mean age years | 84.4 | 84.4 |
| SOFA median (95% CI) | 8 (8-9) | 6 (6-7) |
| CFS<sup>a</sup> median (95% CI) | 4 (4-5) | 4 (4-5) |
| IQCODE<sup>b</sup> median (95% CI) | 3.31 (3.31-3.38) | 3.19 (3.19-3.25) |

<sup>a</sup> CFS = Clinical Frailty Scale; <sup>b</sup> IQCODE=Informant Questionnaire on Cognitive Decline in the Elderly; <sup>c</sup> Norway, Sweden, Poland, Greece, Libya, Austria and Netherland (partially); <sup>d</sup> Denmark, Ireland, Wales, Germany, Belgium, France, Switzerland, Spain, Italy, Portugal, Ukraine, Russia, Turkey, Croatia, Netherlands (partially)

The groups were comparable at admission with regards to age and CFS. SOFA score was higher in group A and the cognitive decline score, IQCODE, was also increased, suggesting worse cognition in this group. With regards to outcomes, the groups differ substantially. Patients in group A with higher degree of organ dysfunction were more often mechanically ventilated and given vasoactive drugs at day 1 and 30-day survival was 50.8% compared to 66.7% in group B (p<0.001) (table 2). No clinical differences in end-of-life decisions between the groups was revealed (data not shown). In the sensitivity analysis, we included England sequentially to either group A and B which did not influence the main results as described above (table 3a and b). Figure 1 reveals the difference in 30-day mortality over the spectrum of SOFA scores in the two groups. In the regression analysis consent or no consent was an independent variable in addition to SOFA score and IQCODE.

**Table 2.** Outcome data in all included patients.

|  | Group A (no consent)<br>n=1220 | Group B (consent) n= 1878 |
| --- | --- | --- |
| LOS hours median (95% CI) | 93 (84-101) | 94 (89-96) |
| Ventilated day 1 (%)<sup>1</sup> | 653 (54%) | 768 (41%) |
| Vasoactive drugs day 1 (%)<sup>1</sup> | 780 (65%) | 844 (45%) |
| 30-day survival (%) <sup>1</sup> | 620 (50.8%; 48.0–<br>53.6%) | 1253 (66.7%; 64.6 – 68.8%) |

<sup>1</sup> p<0.0001 Chi-squared; LOS = length of stay

**Table 3a.**
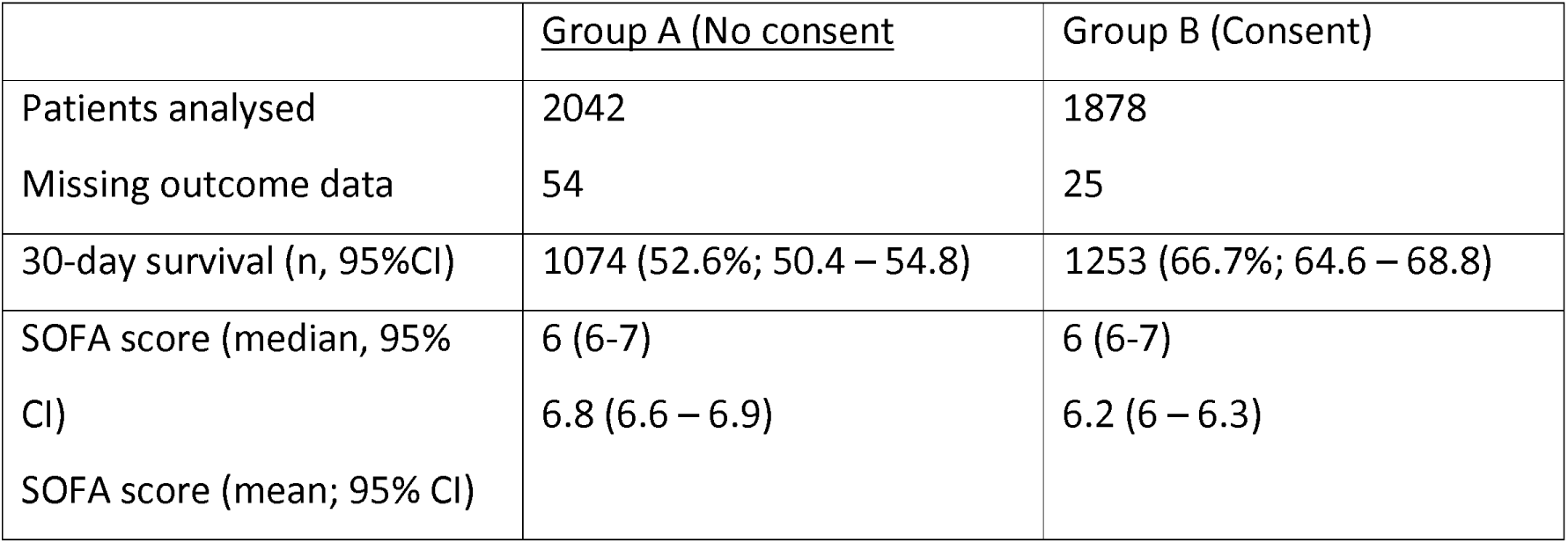
Sensitivity analysis including England added in group A (no consent)

**Table 3b.**
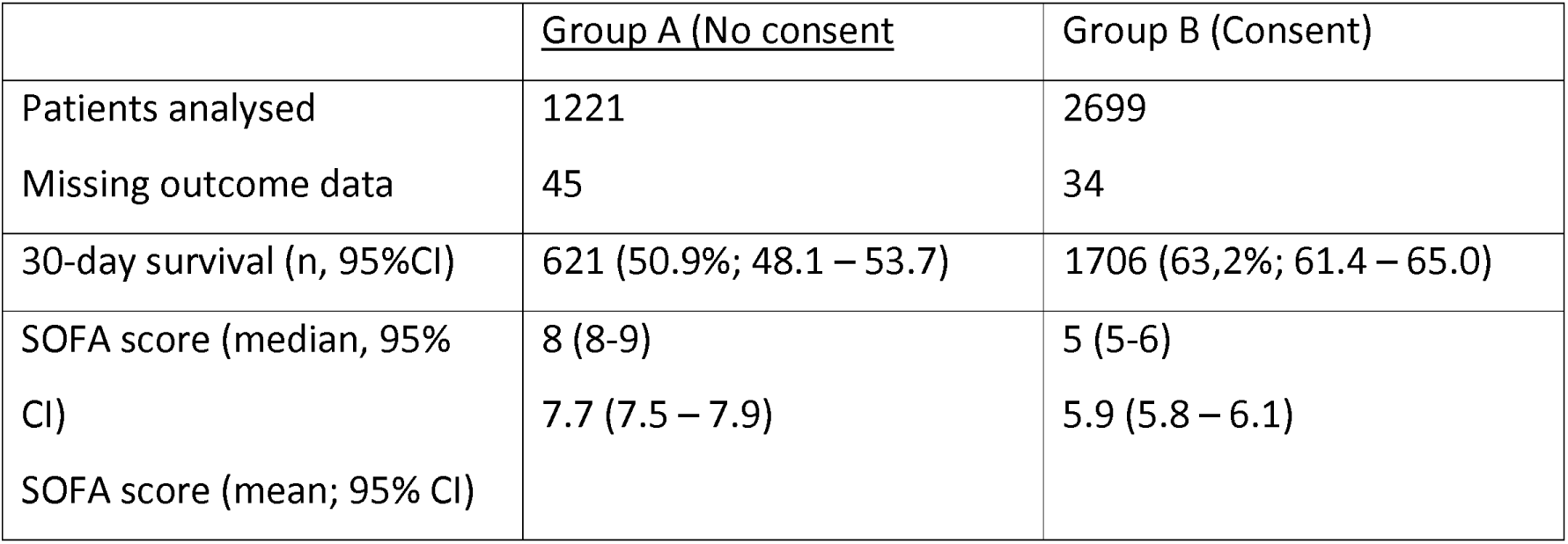
Sensitivity analysis including England in added in group B (consent)

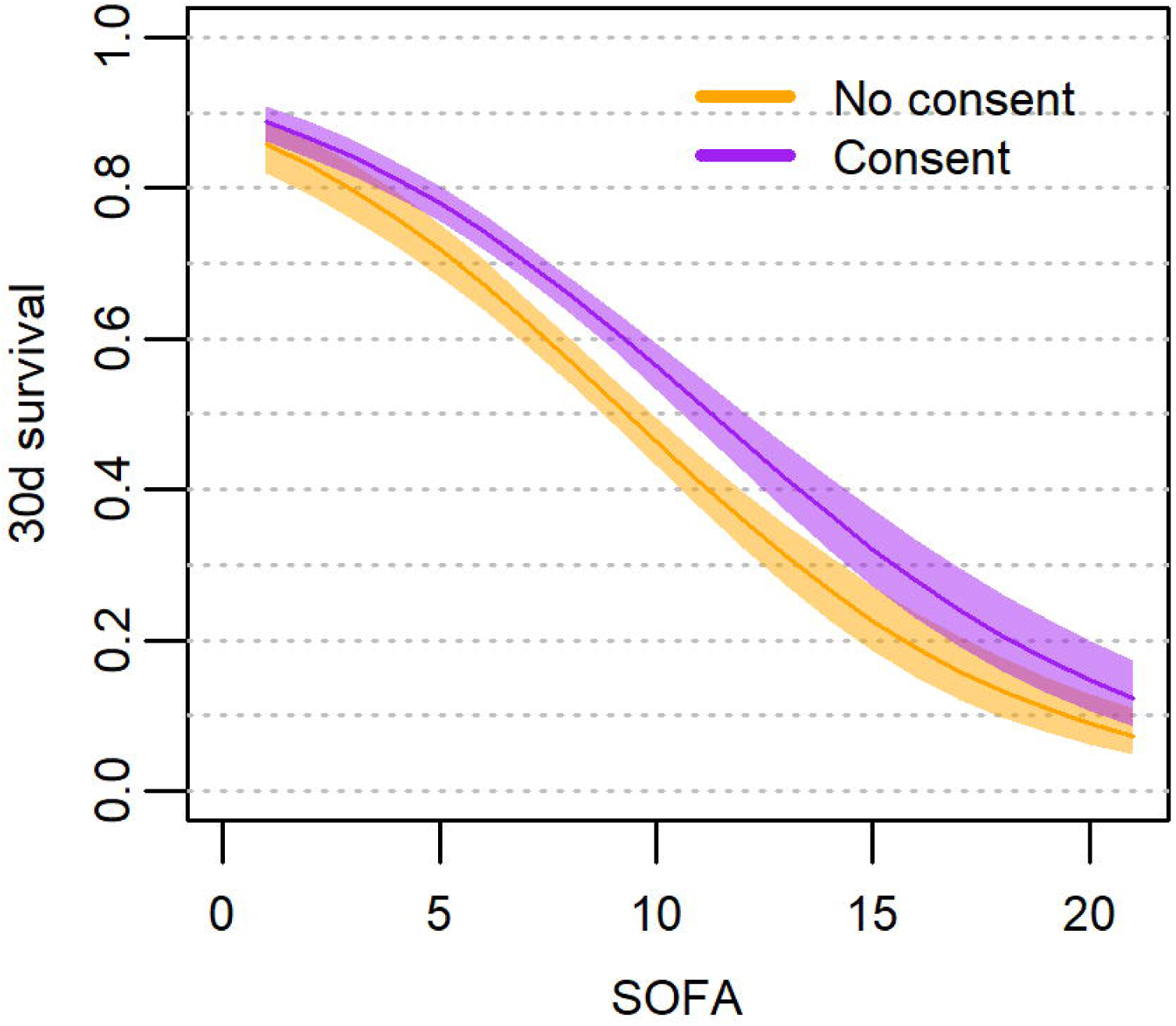

All participating countries obtained ethical consent for conducting this study, although as the study shows the consent to participate varies from country to country. In Norway where the principal investigator (HF) works, the reference from the National Regional Board in Helse Sør-Øst is: 2018/87 (www.etikkom.no).

## Discussion

In this secondary analysis of data from the VIP2 study, we found large differences in the patient cohorts and outcomes when countries were compared with versus without required upfront consent for study inclusion. In a “geriatric” sense they are similar regarding age, frailty, activity of daily life and comorbidities, but considerably different regarding their acute disease severity as estimated by the SOFA score, cognition and the use of life sustaining support on day one. As a consequence, the outcome in the two groups is strikingly different, with 51% survivors when no informed consent was requested necessary compared to 67% in the competing group, an absolute difference of 16%.

One likely explanation for this finding is difficulties in recruiting the most unstable patients when informed consent was required at ICU admission. This seems logical for several reasons. When a critically ill patient is admitted to the ICU, the focus is on the acute treatment and stabilization of the disease process. Confronting patients or caregivers/family in this phase with information and explanation about a clinical study, although with no intervention, often has low priority. If directly asked, many patients are unconscious or with severely reduced mental capacity and caregivers are often too stressed to digest and understand information when asked for surrogate consent. Additionally, in many tertiary centers, family and caregivers may arrive in the ICU after the inclusion window, precluding participation in the study. Hence, sometimes the simplest way to respond is not to give consent. However, this does not mean that such an ICU has equally sick patients, but for the abovementioned reasons, they are just not recruited into the study, resulting in a selection bias of less severely ill patients.

As we have demonstrated, the way consent is handled may be an important confounding factor with considerable implication for the composition of the recruited cohort and hence affecting the primary study endpoint. If the most critically ill unstable patients are left out from inclusion and hence analysis this may give a false impression of a better outcome.

Consent has been the focus in several publications [3], but not discussed in studies involving critically ill old patients. In a study about consent rates documented in clinical studies involving critically ill patients, the researchers found that, particularly in non-randomized studies, information on how consent was handled was lacking in most studies (81%). This was considered a potential source of bias and validity of the studies [4]. The use of surrogates for informed consent in patients on mechanical ventilation has recently been discussed and was associated with difficulties and was not always consistent with the patients’ view [5]. In an interesting discussion about implication of the Food and Drug Administration guideline for informed consent, the authors conclude that inclusion without patient consent should be feasible with the current guideline when patients can not consent [6].

In some countries, like in Norway, there is a special document dealing with inclusion of patients in studies concerning emergency medical conditions [7]. Research, even intervention trials, may be conducted if: 1. The patient is unable to consent (unconscious, unable to comprehend information). 2. Similar research cannot be done in non-emergency situations; and that the project has been approved from the independent local ethical committee. 3. Research should be beneficial for the patient or should be of potential benefit for others and impose minimal extra risks for included patients. 4. If the patient recovers, he/she must be given information about the study at that time and the option to consent or not (deferred consent). However, as our study reveals, in most European countries observational research cannot be performed today without written informed consent at admission. The fact that this is obstructing observational research has been discussed previously [8].

Our study was a purely observational study with no interventions outside standardizing information about geriatric syndromes. Although not potentially beneficial or detrimental for the individual patient, our study’s information may have huge implication for the group of very old patients who are acutely admitted to the ICU. If, as our results may indicate, survival in unselected patients is nearly halved within 30 days, we have the obligation to find ways to reduce unnecessary and futile therapy that is a huge burden on patients as well as care-givers, and to provide such care to those that most likely will profit from such treatment. Moreover, if the external validity of future observational studies is so low that we cannot trust such information, then our research becomes useless or even dangerous.

This study has its strength being a prospective study with a high number of patients from many different countries in Europe, and patients were followed for 30 days. However, a weakness is the post-hoc design of this sub-study, which was not planned for during the pre-study phase. Another weakness is that in some countries with no national guidelines for research Ethical boards, the rules for inclusion could vary from region to region, and this variation have not been possible to capture in this analysis.

## Conclusions

In this study we document the effects on how to receive informed consent in two patient groups within the same prospective study. If consent was necessary at admission (compared to deferred consent patients), patients were less severely ill and with a higher 30-day survival.

We hope that the EU will engage in the harmonization of inclusion rules in clinical scenarios where patients are unable to understand and/or make their own judgment with regard to study participation. The unfortunate alternative would be significant biased data obtained from critically ill patients in Europe.

## Data Availability

All data produced in the present study are available upon reasonable request to the authors

